# Brain-wide Disruptions of Anatomical Connectivity in Antipsychotic-Naïve First Episode Psychosis

**DOI:** 10.1101/2023.11.10.23298391

**Authors:** Sidhant Chopra, Priscila T. Levi, Alexander Holmes, Edwina R. Orchard, Ashlea Segal, Shona M. Francey, Brian O’Donoghue, Vanessa L. Cropley, Barnaby Nelson, Jessica Graham, Lara Baldwin, Hok Pan Yuen, Kelly Allott, Mario Alvarez-Jimenez, Susy Harrigan, Christos Pantelis, Stephen J Wood, Patrick McGorry, Alex Fornito

## Abstract

**OBJECTIVE:** Disruptions of axonal connectivity are thought to be a core pathophysiological feature of psychotic illness, but whether they are present early in the illness, prior to antipsychotic exposure, and whether they can predict clinical outcome remains unknown.

**METHODS:** We acquired diffusion-weighted MRI to map axonal connectivity between each pair of 319 parcellated brain regions in 61 antipsychotic-naive individuals with First Episode Psychosis (FEP; 15-25 years, 46% female) and a demographically matched sample of 27 control participants, along with clinical follow-up data in patients 3 months and 12 months after the scan. We used connectome-wide analyses to map disruptions of inter-regional pairwise connectivity coupled with connectome-based predictive modelling to predict longitudinal change in symptoms and functioning.

**RESULTS:** Individuals with FEP showed disrupted connectivity in a brain-wide network linking all brain regions when compared with controls (*p*_*FWE*_ *=* .03). Baseline structural connectivity significantly predicted change in functioning over 12 months (*r =* .44; *p*_*FWE*_ *=* .041), such that lower connectivity within fronto-striato-thalamic systems predicted worse functional outcomes.

**CONCLUSIONS:** Brain-wide reductions of structural connectivity exist during the early stages of psychotic illness and cannot be attributed to antipsychotic medication. Moreover, baseline measures of structural connectivity can predict change in patient functional outcomes up to one year after engagement with treatment services.

---

The brain is often modelled as a collection of specialized regions connected by a complex web of axonal fibres (Bullmore & Sporns, 2009; Fornito et al., 2016). These fibres enable the coordination of neuronal dynamics and the transport of trophic and other molecules throughout the brain. Since the time of Wernicke, researchers have viewed the symptoms of psychosis as resulting from a disruption of this intricate network of connectivity (Fornito et al., 2012; Friston, 1998; Stephan et al., 2009; Wernicke, 1906), a view supported by a large body of magnetic resonance imaging (MRI) research identifying widespread alterations of axonal connectivity across different stages of psychotic illness (Fornito et al., 2012; Karlsgodt, 2020; Kubicki et al., 2005; Parnanzone et al., 2017).

Recent advances in connectomics have enabled brain-wide mapping of structural dysconnectivity (Fornito et al., 2015; Sporns, 2011). These methods have identified diffuse white-matter pathology in people diagnosed with psychotic illness (Klauser et al., 2017; Zalesky et al., 2011), while other studies have emphasised the role of specific circuits (Van Den Heuvel et al., 2019), including frontotemporal fibre bundles (Voineskos et al., 2010; Wheeler & Voineskos, 2014). The majority of studies to date have been conducted in samples of individuals with established schizophrenia who are taking antipsychotic medication, preventing an understanding of structural alterations proximal to illness onset.

Studies in non-human primates have demonstrated reduced white-matter volume with increasing exposure to antipsychotic medications (Dorph-Petersen et al., 2005). Similarly, longitudinal observational studies in humans examining brain-wide white-matter integrity have consistently demonstrated alterations that coincide with antipsychotic initiation (Meng et al., 2018; Szeszko et al., 2014). Together, these studies suggest that antipsychotic exposure may be masking, or be mistaken for, illness-related structural dysconnectivity. The few previous studies examining whole-brain structural connectivity in antipsychotic-naïve first-episode samples have focused on topological or global measures of connectivity, reporting less effective organizational patterns and overall decreases in connectivity (Li et al., 2018; Zhang et al., 2015). However, the specific connections affected, and their clinical relevance is unknown. Moreover, most studies in patients have focused on how structural alterations are associated with current symptoms assessed cross-sectionally rather than on predicting how clinical outcomes change longitudinally. Such longitudinal changes are more clinically relevant, having implications for early detection and intervention strategies, but no prior study has examined whether structural connectivity can predict longitudinal changes in symptoms or other outcomes at the level of individual antipsychotic-naïve patients. The clinical relevance of structural dysconnectivity in psychosis is thus unclear.

Here, we aimed to test whether alterations in structural connectivity are isolated to specific circuits or diffusely distributed throughout the brain within antipsychotic-naïve individuals experiencing a first-episode of psychosis. Our primary aims were to (1) comprehensively map alterations in structural connectivity during the initial stages of psychotic illness; and (2) examine whether structural connectivity can predict longitudinal changes in psychiatric symptoms and functional outcomes at three- and twelve-months following engagement with treatment services.

## Method

### Study Design

The study took place at the Early Psychosis Prevention and Intervention Centre, which is part of Orygen Youth Health, Melbourne, Australia. The current study uses data from a larger trial registered with the Australian New Zealand Clinical Trials Registry in November 2007 (ACTRN12607000608460) and received ethics approval from the Melbourne Health Human Research and Ethics committee. After entering the study at baseline, patients were randomized to one of two groups: one given antipsychotic medication plus intensive psychosocial therapy and the other given a placebo plus intensive psychosocial therapy. A demographically matched healthy control group with no psychiatric diagnoses was also recruited. For both patient groups, the treatment period spanned six months. Diffusion-weighted Imaging (DWI), structural T1w, and functional MRI scans were acquired at baseline, prior to randomization into a treatment arm. Structural T1w, functional MRI, and clinical assessments were also conducted at baseline, 3 and 12 months, and have been analysed in other reports (Chopra et al., 2020; Chopra et al., 2021; Chopra et al., 2023). Here, we focus on the diffusion MRI data, which were only acquired at baseline. The randomization phase of the study terminated at 6 months, so patients in either group could have received antipsychotic medication and ongoing psychosocial interventions between 6 and 12 months into the study. Further research and safety protocols can be found elsewhere (Francey, O’Donoghue, et al., 2020; O’Donoghue et al., 2019).

### Participants

We recruited 61 people aged 15-25 years (46% female) who were experiencing a first episode of psychosis, defined as fulfilling Structured Clinical Interview for DSM-IV criteria for a psychotic disorder (see Supplementary Table 1 for details). We repeated primary analyses after excluding individuals with a missing diagnosis or cannabis-induced psychotic disorder (see Results). We also recruited a demographically matched control group (n=27) without a psychotic diagnosis. Additional patient inclusion criteria to minimise risk were: ability to provide informed consent; comprehension of English language; no contraindication to MRI scanning; duration of untreated psychosis of less than 6 months; living in stable accommodation; low risk to self or others; none or minimal previous exposure to antipsychotic medication (< 7 days of use or lifetime 1750mg chlorpromazine equivalent exposure; further details provided in Supplementary table 1).

### MRI acquisition and processing

Diffusion-weighted imaging (DWI) and structural T1-weighted data were acquired using a 3T Siemens Trio Tim scanner at the Royal Children’s Hospital in Melbourne, Australia. Acquisition parameters are provided in the Supplement. Whole-brain probabilistic tractography was preformed was performed using standardized, reproducible workflows (Al-Sharif et al., 2020; Theaud et al., 2020). Importantly, probabilistic tractography algorithms are prone to false positives and do not directly index the quantitative strength of connections between pairs of regions (Maier-Hein et al., 2017; Schilling et al., 2019). We therefore implemented a state-of-the-art optimisation procedure, Convex Optimization Modelling for Microstructure Informed Tractography (COMMIT2), which is superior to other methods on key benchmarks derived from fibre-tracking phantoms (Nelson et al., 2023; Schiavi et al., 2020). COMMIT2 filters and re-weights pair-wise connections strengths according to their contribution to the observed DWI signal, providing more biologically accurate and quantitative estimates of connectivity than traditional streamline count measures. To create whole-brain structural connectivity matrices, COMMIT2-re-weighted streamlines were assigned to the closest region, as defined by previously validated cortical (Schaefer et al., 2018) and subcortical (Tian et al., 2020) atlases, yielding undirected structural connectivity matrices defining pair-wise connections for 319 parcellated regions. A detailed overview of DWI processing and optimisation can be found in the Supplement.

### Statistical analysis

#### Group-level differences in structural connectivity

Most studies of structural connectivity rely on standard statistics to compare group mean values of connectivity estimates (e.g., *t*-tests, linear regressions). This approach ignores the fact that such estimates often have a highly non-Gaussian distribution because many participants may not have a reconstructed connection linking a given pair of regions, meaning that a proportion of individuals will have a connectivity weight of zero while the remaining participants have some non-zero weight estimate.

To address this problem, we fitted zero-inflated gamma regression models at each pairwise connection (Mills, 2013). The zero-inflated gamma regression model is a two-part hurdle model that encompasses a zero-inflation component and a gamma component. The zero-inflation component, modelled using a logistic regression, accounts for the difference in the probability of observing a zero at a given edge. The gamma component, using a logarithmic link function, characterizes the differences in positive non-zero connectivity strength at a given edge. All models were adjusted for age, sex, and mean framewise displacement (head motion). Detailed information regarding the zero-inflated gamma regression model can be found in the Supplement.

The Network Based Statistic (NBS) was used to perform familywise error-corrected (FWE) inference at the level of connected components of edges (Zalesky et al., 2010), with the primary component-forming threshold, τ, set to *p* < .05. Further statistical details can be found in the Supplement as well as results obtained with τ *=* .01 and τ *=* .001 (SFig1).

To comprehensively delineate changes in structural connectivity across the 21,476 different connections, we present the results at three different scales: (1) the individual edge level, embedded in the spatial layout of the brain (e.g., Fig 1A); (2) the level of individual brain regions, to identify specific brain areas attached to a high number of connections implicated in the NBS subnetwork (e.g., Fig1F/L); and (3) the network-level, in which different regions are aggregated in one of 10 canonical brain networks (Schaefer et al., 2018; Tian et al., 2020; Yeo et al., 2011) where we show the proportions of affected edges both within and between these networks (e.g., Fig1B-C).

**Figure 1.**
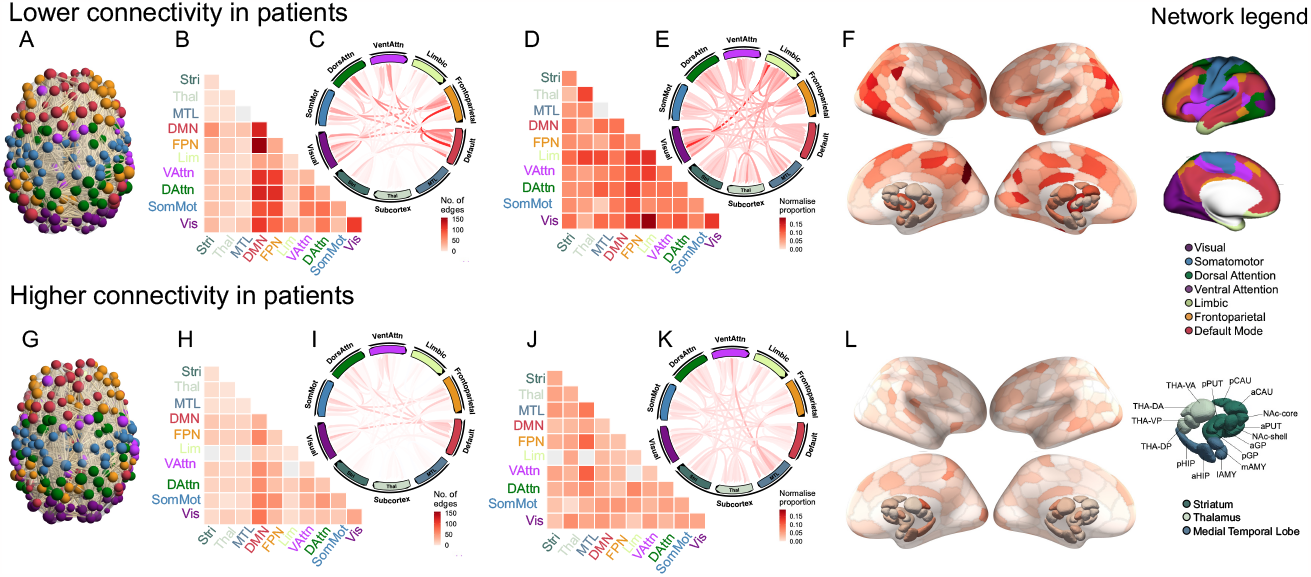
Top row (**A-F**) shows lower connectivity in patients and the bottom row (**G-L**) shows higher connectivity in patients. **A/G)** Visualisation of the significant NBS subnetwork, with the nodes coloured by network and weighted by degree; **B/H)** heatmap of the proportion of edges within the NBS component that fall within each of the canonical networks as represented quantified using raw and **D/H**) normalized proportions (see Methods for details); **C/E-I/K)** a circle plot of the proportion of edges within the NBS component that fall within each of the canonical networks as represented quantified using raw (**C/I**) and normalized **(E/K**) proportion; **F/L**) surface renderings depicting the number of edges in the NBS subnetwork attached to each brain region. Legend shows Schaefer 300 region (Schaefer et al., 2018) and Tian subcortex 32 region (Tian et al., 2020) atlas. a Indicates anterior; AMY, amygdala; CAU, caudate nucleus; d, dorsal; DA, dorsoanterior; DMN, default mode network; DAttn, dorsal attention network; DP, dorsoposterior; FPN, frontoparietal/control network; GP, globus pallidus; HIP, hippocampus; l, lateral; Lim, cortical limbic network; m, medial; MTL, medial-temporal lobe (amygdala and hippocampus); NAc, nucleus accumbens; p, posterior; SomMot, somatomotor network; Stri, striatum; PUT, putamen; THA, thalamus

To determine whether the observed structural connectivity alterations showed any network-specificity, we calculated the proportion of edges within a given NBS component that fell within each of ten canonical brain networks (e.g., Fig1B – upper triangle of matrix). Different brain networks have intrinsic differences in size, meaning that larger networks will generally have a higher likelihood of being implicated. We therefore present both raw proportions and proportions normalized by the total number of possible network connections (edges included in the final analysis) between each pair of networks (e.g., Fig1B – lower triangle of matrix); the former identifies preferential involvement of a given network in an absolute sense while the latter accounts for differences in the number of potential connections (i.e., the tendency for highly interconnected networks to be more likely to be implicated in a given NBS network).

#### Connectome-based prediction of clinical outcomes

We used connectome based predictive modelling (CPM; Shen et al., 2017) to assess whether structural connectivity at baseline can predict longitudinal change in patient symptoms and functioning from baseline to 3-months (Δ3-months) and baseline to 12-months (Δ12-months). We computed proportional change scores (*y*_1_ − *y*_2_/*y*_1_, where *y*_1_ and *y*_2_ correspond to the clinical scale values measured at time 1 and 2, respectively) for the two preregistered primary trial outcome measures (Francey, O’Donoghue, et al., 2020)––total scores for the Social and Occupational Functioning Assessment Scale (SOFAS) and the Brief Psychiatric Assessment Scale (BPRS)––at each of two longitudinal timepoints (Δ3-months, Δ12-months). For Δ3-months and Δ12-months analysis, 34 and 31 patients with complete data at both timepoints, respectively, were included in the analysis. Change score on both measures did not significantly differ between the two treatment groups at Δ3-months or Δ12-months (*all p* > .29). Distributions of change scores for each scale at each timepoint are provided in SFig2.

We implemented CPM using nested *k* -fold cross-validation. First, the structural connectivity weights and behavioural change scores were divided into four distinct subsets or ‘folds’ of subjects. The model was trained on three folds, while the fourth, or ‘held out’, fold remained separate. The training process involved iteratively selecting different sets of three folds for training and using the model to predict outcomes for the fourth fold. This was repeated four times, with each fold being held out once. To train the model, a Spearman correlation between structural connectivity and the behavioural change score was computed within the training set and edges with *p* < .01 uncorrected were selected as features (Shen et al., 2017). To ensure robustness of our findings, significant models were repeated using *p* < .05 and *p* < .001 as alternate feature selection thresholds (see SFig4). Then for each subject, the structural connectivity weights of the selected positively and negatively correlated edges were summed separately, resulting in two summary metrics per subject. These metrics were used to train two separate general linear models to predict the behavioural change score, and to predict the behavioural change score in the held-out test set. The product-moment correlation between observed and predicted change score within the held-out test set was used to assess prediction performance.

The entire *k* -fold modelling process was repeated 100 times to estimate the sampling error of the model’s prediction performance. To assess model significance, we permuted the behaviour scores 1000 times and repeated the k-fold modelling process, resulting in a null distribution of product-moment correlations. We then estimated *p*-values as the proportion of null correlations greater than the mean observed model performance across the 100 repeated runs of the *k*-fold modelling process (*r*_*mean*_).

Overall, eight primary prediction models were run, corresponding to two primary behavioural change scores (SOFAS, BPRS) at two timepoints (Δ3-months, Δ12-months), using two separate feature selection methods (edges positively or negatively correlated with the outcome measures). For each model, the null permutations began with the same computational seed, ensuring that the same permutation scheme was used at each null across models. To implement FWE correction across the eight primary models, we retained the maximum product-moment correlation across each of these models for each of 1000 permutations. This allowed us to compute a FWE corrected *p*-value for each model (Alberton et al., 2020; Winkler et al., 2014), with statistical significance assessed at *p*_*FWE*_ < .05. In addition to these models examining primary outcomes, we repeated the above process using four exploratory secondary outcome measures: Scale for the Assessment of Negative Symptoms (SANS), Hamilton Anxiety Rating Scale (HAMA), Hamilton Depression Rating Scale (HAMD) and World Health Organization Quality of Life Scale (WHOQOL).

To assess which edges were the most strongly implicated in predicting longitudinal changes in behaviour within significant models, we examined the network- and region-level architecture of edges that were selected as part of the feature selection process in at least half (50%) of the 100 iterations of k-fold cross-validation. Interpretations based on more conservative thresholds of 80% and 100% are provided in the supplement (SFig5). All code used for MRI processing, statistical analyses and generating figures can be found online at https://github.com/sidchop/PsychosisConnectome.

## Results

### Demographics and clinical characteristics

We have previously reported the demographics and clinical characteristics of this cohort. Briefly, patient and control samples did not differ significantly in sex or handedness, but the patients were, on average, 1.9 years younger and had 2 years less education (see Supplement for further details).

### Connectome-wide disruptions of structural connectively in antipsychotic-naïve FEP

We identified a single NBS component showing widespread structural dysconnectivity in patients compared with controls, comprising 3209 edges (14.9%) linking all 319 regions (*p*_*FWE*_ *=* .03; Figure 1A). Within this network, 2681 edges (68.7%) showed lower connectivity and 1221 edges (31.3%) showed higher structural connectivity in patients.

Evaluating raw proportions, connections associated with reduced structural connectivity in patients were predominantly concentrated in the default mode and frontoparietal networks (Fig1B). Evaluating normalized proportions, which emphasize network involvement after accounting for differences in the total number of possible connections, the disruptions showed a more homogenous distribution, implicating a similar proportion of connections in each network (Fig1B, Fig1D). At a regional level, right precuneus, parietal cortex, left temporal pole, visual cortex, and posterior putamen were among the areas most strongly implicated in the network of lower structural connectivity (Fig1D).

Considering connections showing increased structural connectivity in patients, raw proportions indicated a predominant concentration within the default mode network (Fig1G, Fig1H), but once again normalized proportions pointed to a more homogeneous distribution across networks. At a regional level, the right dorsal anterior thalamus, posterior cingulate, visual and somatomotor cortices were among the areas most strongly implicated in the network of higher structural connectivity.

These findings remained largely consistent when excluding individuals with missing or substance-induced psychosis diagnoses (SFig1), and when using different zero-inflation thresholds for determining the minimum number of zero values within an edge prior to exclusion (SFig6). The findings also remained consistent using primary component-forming thresholds of τ *=* 0.01, but not τ *=* 0.001, suggesting that the detected effect is diffuse and spatially widespread (SFig3). We did not detect any significant differences in connectivity between the two treatment groups at baseline.

### Structural connectivity reliably predicts longitudinal changes in functioning

Structural connectivity significantly predicted change in functioning (SOFAS) over 12-months (*r*_*mean*_ *=* .44; *p*_*FWE*_ *=* .041; Figure 2A). This model considered edges that were positively correlated with behavioural change as features, indicating that lower axonal connectivity was robustly associated with lower functional recovery during the early stages of psychosis. A large proportion of the most reliably predictive structural connections were found between the default mode and frontoparietal networks (Fig2B-C). After accounting for network size, connections within limbic and thalamic regions, and between striatum and limbic regions, were preferentially implicated (Fig2C). At a regional level, bilateral medial and lateral prefrontal cortex and posterior putamen were among the areas most strongly contributing to prediction (Fig2D). No significant associations were detected for symptoms (BPRS) or for change scores over 3-months.

**Figure 2.**
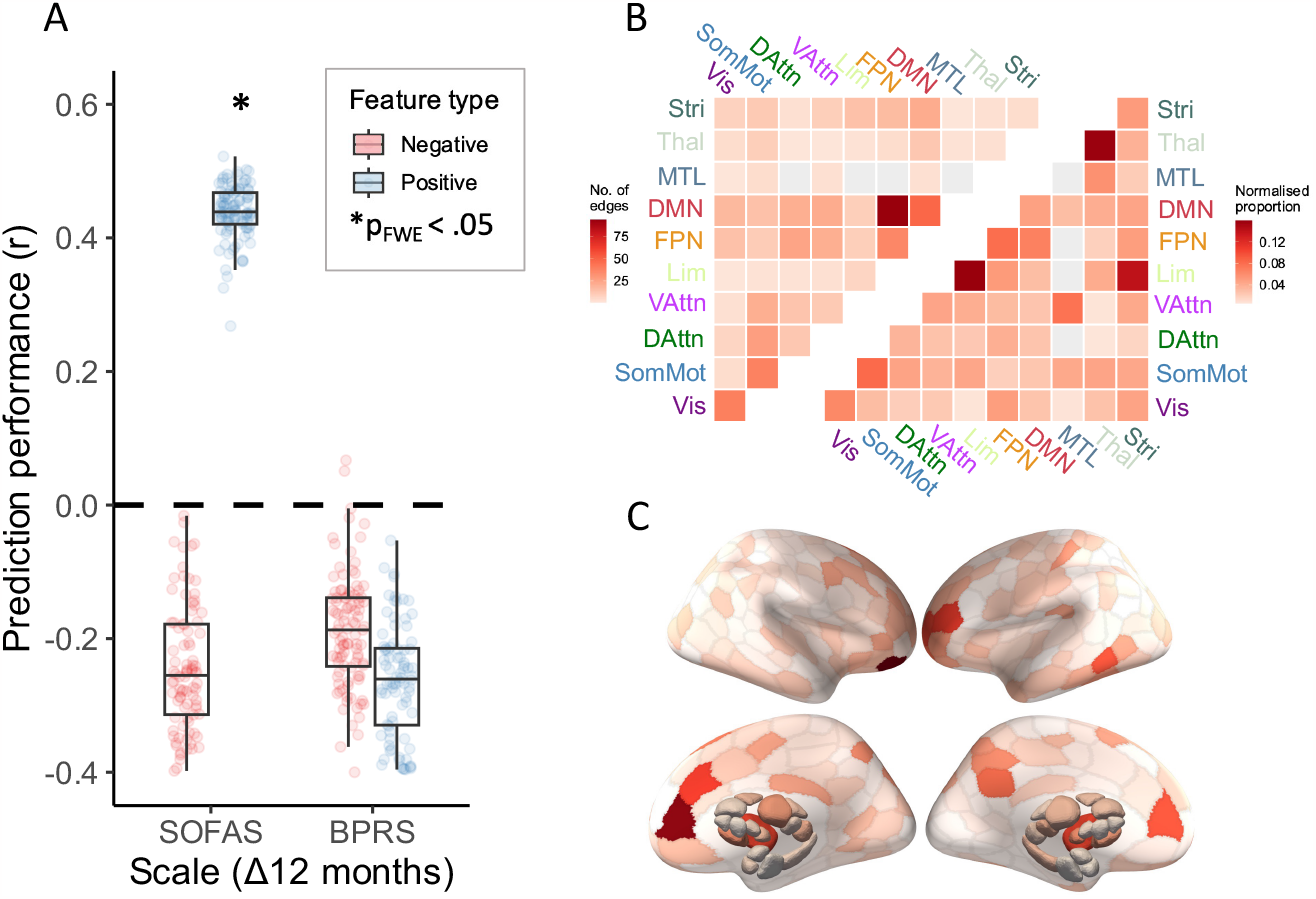
**A)** Product-moment correlation between observed and predicted behavioral change score (prediction performance) between baseline and 12-months for the two primary trial outcome measures (Social and Occupational Functional Assessment Scale; SOFAS; Brief Psychiatric Rating Scale; BPRS). For each of the scales, two models were computed, that selected connections that were either positively (blue) or negatively (red) correlated with the outcome measure. **B)** A heatmap of the proportion of edges robustly implicated in predicting change in SOFAS (higher connectivity predicts improvement) within the prediction subnetwork that fall within each of the canonical networks as represented quantified using raw (upper triangle) and normalized (lower triangle) proportions (see Methods for details). **C)** surface renderings depicting the number of edges in the prediction subnetwork attached to each brain region.

For the secondary outcome measures, structural connectivity again significantly predicted change in Δ12-months functioning, this time using the WHOQ scale (*r*_*mean*_ *=* .41; *p =* .013), but this effect did not survive FWE correction. Similar regional and network level connections were implicated as in the primary SOFAS prediction model (SFig7). Prediction performance for all models is provided in the SFig8.

To assess the impact of potential confounders, we examined correlations between behavioural change scores on the two primary outcome measures (SOFAS and BPRS) and age, sex, head motion, antipsychotic medication exposure (total cumulative olanzapine equivalent dose). No significant correlations were detected between behavioural change scores and these variables for either baseline to 3-month or baseline to 12-month time points (all *p* > .05).

## Discussion

Whether disruptions of structural connectivity in people with psychosis are concentrated within specific circuits or diffusely distributed across the entire brain, and whether they are attributable to antipsychotic medication, or clinically relevant, has thus far remained unclear. We used advanced whole-brain tractography to map differences in structural connectivity between antipsychotic-naïve individuals experiencing a first episode of psychosis and a normative control group, finding a diffuse, brain-wide network of reduced connectivity that was evenly distributed across all canonical brain networks. We further found that structural connectivity can robustly predict changes in general functioning at the individual patient-level, with lower connectivity within fronto-striato-thalamic circuits predicting worse outcomes over one year.

### Wide-spread differences in structural connectivity in antipsychotic-naïve psychotic illness

Fibre bundles largely showed lower connectivity in patients compared to healthy controls and implicated an extensive network connecting all brain regions. This result aligns with previous research showing that hypoconnectivity is far more widespread than previously indicated by studies examining isolated regions (Klauser et al., 2017; Repple et al., 2023; Zalesky et al., 2011). Our findings further extend this research by demonstrating that these alterations are present even in the very early stages of illness onset and in the absence of antipsychotic medication exposure, suggesting that they are directly associated with illness-related processes.

Some studies have revealed more severe and extensive structural connectivity changes in later stages of illness, suggesting potentially progressive white matter pathology (Cetin-Karayumak et al., 2020; Cropley et al., 2017). These previous findings are based on observations of subtle or spatially confined alterations observed in the early stages of illness, compared to later stages (Di Biase et al., 2017). In contrast, our findings demonstrate that prominent and widespread dysconnectivity is present at least from the outset of psychosis. This discrepancy could be attributed to the increased sensitivity of our current study due to the application of state-of-the-art streamline filtering and reweighting, resulting in a sensitive and biologically interpretable indicator of structural connectivity (Nelson et al., 2023; Schiavi et al., 2020). Additionally, we employed zero-inflated generalised linear models to address both zero-inflation and non-Gaussian distributions in brain-wide mass-univariate analyses, negating the need for the arbitrary exclusion of large proportions of connections. These considerations have been largely overlooked in previous investigations of whole-brain structural connectivity and have potentially resulted in lower sensitivity to detect connectivity alterations.

Since our findings show dysconnectivity prior to starting antipsychotic treatment, it is possible that later-stage connectivity changes, to the extent that they progress, may result from prolonged antipsychotic use. Indeed, the only two blinded, randomised and placebo-controlled studies of antipsychotic-naïve individuals with psychosis have demonstrated a decline in brain-wide white-matter integrity (Voineskos et al., 2020) and cerebellar white-matter volume (Chopra et al., 2020) following antipsychotic treatment, with the latter study conducted in the same cohort of patients used here. Thus, while the early-stage connectivity alterations detected in the current study could represent initial stages of progressive pathology, longitudinal studies examining whole brain connectivity across both prodromal and latter stages of illness would be required to test this hypothesis. Indeed, an alternative hypothesis is that our findings could represent altered neurodevelopment. Prior longitudinal work has shown that white matter alterations may not worsen during transition to illness, but may represent an altered neurodevelopmental course marked by early maturational peaks and premature white matter decline (Di Biase et al., 2021) or accelerated age-related decline (Cropley et al., 2017).

While we find that the largest number of affected connections fall between the default mode and frontoparietal network and the rest of the brain, when normalizing for network size we find a substantially more homogenous distribution of affected networks. Therefore, the preferential implication of specific systems may largely be a function of the larger number of regions comprising higher-order association networks. Our findings align with previous work showing spatially diffuse intra-axonal connectivity, as opposed to localised alterations in white matter (Klauser et al., 2017). Although the precise causal mechanisms underlying diffuse dysconnectivity remain unknown, several factors may contribute, including genes (Anderson et al., 2020; Bohlken et al., 2016; Romme et al., 2017), inflammation (Trépanier et al., 2016; Wang et al., 2020), and social determinants such as low socioeconomic status (Qiu & Liu, 2023; Rakesh & Whittle, 2021), or lifestyle factors such as poor nutrition, physical inactivity, and substance use (Moser et al., 2018; Raghava et al., 2021).

### Structural connectivity predicts longitudinal changes in functioning

Our prediction modelling revealed that lower structural connectivity within fronto-striato-thalamic circuits at baseline can robustly predict declines in functional outcome up to 12-months post-baseline. To our knowledge, our findings provide the first evidence demonstrating that axonal connectivity in these circuits has ramifications for functional recover in FEP. Alterations of fronto-striato-thalamic circuits have long been implicated in the pathogenesis of psychosis, with functional alterations present across all stages of psychosis (Dandash et al., 2014; Dandash et al., 2017; Fornito et al., 2013; Sabaroedin et al., 2023; Sabaroedin et al., 2022; Sabaroedin et al., 2019) and structural alterations reported in first episode (Quan et al., 2013), first-degree relatives (De Leeuw et al., 2015), and established schizophrenia (Levitt et al., 2017; Zhao et al., 2020). Treatment response in first-episode patients is associated with increased functional coupling in these same circuits (Chopra et al., 2021; Sarpal et al., 2016; Sarpal et al., 2015) as well as global structural connectivity patterns (Crossley et al., 2017). Our findings thus suggest that lower inter-regional axonal connectivity within these circuits may be a prospective marker of more severe illness that is less likely to show improvements in functional outcome or potentially respond to treatment. Alternatively, disruptions of these circuits may represent a neurodevelopmental aberration that limits the remediation of circuit functioning, preventing recovery after psychosis onset. Future studies should examine whether the prediction of functional outcomes using the connectivity in these circuits is specific to psychotic illness.

### Limitations and conclusions

The practical challenges of recruiting medication-naïve individuals limited the sample size of the current cohort. It is possible that sample characteristics such as lack of medication exposure or differences in illness severity may impact the generalisability of our findings, although the functional and symptom ratings of our patients are comparable to epidemiologically representative or ‘markedly ill’ first-episode psychosis cohorts (Henry et al., 2010; Leucht et al., 2005; see Chopra et al., 2021 for discussion on generalisability). Our approach is also limited by the accuracy of diffusion MRI (Maier-Hein et al., 2017). While the processing procedures we applied enhanced the biological validity of our connectivity measures as much as possible (Schiavi et al., 2020) further developments in non-invasive connectivity mapping and tractography will be required to allow more precise mechanistic inferences.

In conclusion, diffuse brain-wide reductions of structural connectivity exist during the early stages of psychotic illness and cannot be attributed to antipsychotic medication. Structural connectivity in fronto-striato-thalamic circuits prospectively predicts functional recovery up to one year following engagement with treatment services.

## Data Availability

All data produced in the present study are available upon reasonable request to the authors. Code use for analysis is available online at https://github.com/sidchop/PsychosisConnectome.

## Acknowledgments

This study has been supported by a large number of clinical staff at Orygen Youth Health: Craig Macneil, Kingsley Crisp, Dylan Alexander, Tina Proffitt, Rachel Tindall, Jennifer Hall, Lisa Rumney, Franco Scalzo, Melissa Pane, Linda Kader, Frank Hughes, Clare Shelton, Ryan Kaplan, David Hallford, Bridget Moller, Rick Fraser and research assistants: Daniela Cagliarini, Suzanne Wiltink, Janine Ward and Sumudu Mallawaarachichi. The trial took place at the Early Psychosis Prevention and Intervention Centre, which is part of Orygen Youth Health, Melbourne, Australia. Janssen-Cilag partially supported the early years of this study with an unrestricted investigator-initiated grant and provided risperidone, paliperidone and matched placebo for the first 30 participants. The study was then funded by an Australian National Health and Medical Research Project grant (1064704). The funders had no role in study design, data collection, data analysis, data interpretation, writing, approval or submission of this manuscript.

## References

Al-Sharif, N. B., St-Onge, E., Theaud, G., Evans, A. C., & Descoteaux, M. (2020). Processing the diffusion-weighted magnetic resonance imaging of the PING dataset. bioRxiv, 2020.2011.2024.396549. 10.1101/2020.11.24.396549

Alberton, B. A. V., Nichols, T. E., Gamba, H. R., & Winkler, A. M. (2020, August 1, 2020). Multiple testing correction over contrasts for brain imaging. Neuroimage, 216, 116760. 10.1016/j.neuroimage.2020.116760

Anderson, K. M., Collins, M. A., Chin, R., Ge, T., Rosenberg, M. D., & Holmes, A. J. (2020). Transcriptional and imaging-genetic association of cortical interneurons, brain function, and schizophrenia risk. Nature Communications, 11(1), 2889.

Bohlken, M. M., Brouwer, R. M., Mandl, R. C., Van den Heuvel, M. P., Hedman, A. M., De Hert, M., Cahn, W., Kahn, R. S., & Pol, H. E. H. (2016). Structural brain connectivity as a genetic marker for schizophrenia. JAMA Psychiatry, 73(1), 11–19.

Bullmore, E., & Sporns, O. (2009). Complex brain networks: graph theoretical analysis of structural and functional systems. Nature Reviews Neuroscience, 10(3), 186–198.

Cetin-Karayumak, S., Di Biase, M. A., Chunga, N., Reid, B., Somes, N., Lyall, A. E., Kelly, S., Solgun, B., Pasternak, O., & Vangel, M. (2020). White matter abnormalities across the lifespan of schizophrenia: a harmonized multi-site diffusion MRI study. Molecular Psychiatry, 25(12), 3208–3219.

Chopra, S., Fornito, A., Francey, S., O’Donoghue, B., Cropley, V., Nelson, B., Graham, J., Baldwin, L., Tahtalian, S., Yuen, H. P., Allott, K., Alvarez-Jimenez, M., Harrigan, S., Sabaroedin, K., Pantelis, C., Wood, S. J., & McGorry, P. (2020, January 1, 2020). Differentiating the Effect of Medication and Illness on Brain Volume Reductions in First-Episode Psychosis: A Longitudinal, Randomized, Triple-blind, Placebo-controlled MRI study. medRxiv, 2020.2003.2018.20038471. 10.1101/2020.03.18.20038471

Chopra, S., Francey, S. M., O’Donoghue, B., Sabaroedin, K., Arnatkeviciute, A., Cropley, V., Nelson, B., Graham, J., Baldwin, L., & Tahtalian, S. (2021). Functional connectivity in antipsychotic-treated and antipsychotic-naive patients with first-episode psychosis and low risk of self-harm or aggression: a secondary analysis of a randomized clinical trial. JAMA Psychiatry, 78(9), 994–1004.

Chopra, S., Segal, A., Oldham, S., Holmes, A., Sabaroedin, K., Orchard, E. R., Francey, S. M., O’Donoghue, B., Cropley, V., Nelson, B., Graham, J., Baldwin, L., Tiego, J., Yuen, H. P., Allott, K., Alvarez-Jimenez, M., Harrigan, S., Fulcher, B. D., Aquino, K., Pantelis, C., Wood, S. J., Bellgrove, M., McGorry, P. D., & Fornito, A. (2023). Network-Based Spreading of Gray Matter Changes Across Different Stages of Psychosis. JAMA Psychiatry. 10.1001/jamapsychiatry.2023.3293

Cropley, V. L., Klauser, P., Lenroot, R. K., Bruggemann, J., Sundram, S., Bousman, C., Pereira, A., Di Biase, M. A., Weickert, T. W., Weickert, C. S., Pantelis, C., & Zalesky, A. (2017, Mar 1). Accelerated Gray and White Matter Deterioration With Age in Schizophrenia. Am J Psychiatry, 174(3), 286–295. 10.1176/appi.ajp.2016.16050610

Crossley, N. A., Marques, T. R., Taylor, H., Chaddock, C., Dell’Acqua, F., Reinders, A. A., Mondelli, V., DiForti, M., Simmons, A., & David, A. S. (2017). Connectomic correlates of response to treatment in first-episode psychosis. Brain, 140(2), 487–496.

Dandash, O., Fornito, A., Lee, J., Keefe, R. S. E., Chee, M. W. L., Adcock, R. A., Pantelis, C., Wood, S. J., & Harrison, B. J. (2014, 2014/07/01). Altered Striatal Functional Connectivity in Subjects With an At-Risk Mental State for Psychosis. Schizophrenia Bulletin, 40(4), 904–913. 10.1093/schbul/sbt093

Dandash, O., Pantelis, C., & Fornito, A. (2017, February 1, 2017). Dopamine, fronto-striato-thalamic circuits and risk for psychosis. Schizophrenia Research, 180, 48–57. 10.1016/j.schres.2016.08.020

De Leeuw, M., Bohlken, M. M., Mandl, R. C., Kahn, R. S., & Vink, M. (2015). Reduced fronto–striatal white matter integrity in schizophrenia patients and unaffected siblings: a DTI study. npj Schizophrenia, 1(1), 1–6.

Di Biase, M., Cropley, V., Baune, B., Olver, J., Amminger, G., Phassouliotis, C., Bousman, C., McGorry, P., Everall, I., & Pantelis, C. (2017). White matter connectivity disruptions in early and chronic schizophrenia. Psychological Medicine, 47(16), 2797–2810.

Di Biase, M. A., Cetin-Karayumak, S., Lyall, A. E., Zalesky, A., Cho, K. I. K., Zhang, F., Kubicki, M., Rathi, Y., Lyons, M. G., Bouix, S., Billah, T., Anticevic, A., Schleifer, C., Adkinson, B. D., Ji, J. L., Tamayo, Z., Addington, J., Bearden, C. E., Cornblatt, B. A., Keshavan, M. S., Mathalon, D. H., McGlashan, T. H., Perkins, D. O., Cadenhead, K. S., Tsuang, M. T., Woods, S. W., Stone, W. S., Shenton, M. E., Cannon, T. D., & Pasternak, O. (2021, 2021/05/24). White matter changes in psychosis risk relate to development and are not impacted by the transition to psychosis. Molecular Psychiatry. 10.1038/s41380-021-01128-8

Dorph-Petersen, K.-A., Pierri, J. N., Perel, J. M., Sun, Z., Sampson, A. R., & Lewis, D. A. (2005, 2005-09-09). The Influence of Chronic Exposure to Antipsychotic Medications on Brain Size before and after Tissue Fixation: A Comparison of Haloperidol and Olanzapine in Macaque Monkeys. Neuropsychopharmacology, 30(9), 1649–1661. 10.1038/sj.npp.1300710

Fornito, A., Harrison, B. J., Goodby, E., Dean, A., Ooi, C., Nathan, P. J., Lennox, B. R., Jones, P. B., Suckling, J., & Bullmore, E. T. (2013, 2013/11/01). Functional Dysconnectivity of Corticostriatal Circuitry as a Risk Phenotype for Psychosis. JAMA Psychiatry, 70(11), 1143–1151. 10.1001/jamapsychiatry.2013.1976

Fornito, A., Zalesky, A., & Breakspear, M. (2015, 2015). The connectomics of brain disorders. 10.1038/nrn3901

Fornito, A., Zalesky, A., & Bullmore, E. (2016). Fundamentals of brain network analysis. Academic press.

Fornito, A., Zalesky, A., Pantelis, C., & Bullmore, E. T. (2012, 10/2012). Schizophrenia, neuroimaging and connectomics. Neuroimage, 62(4), 2296–2314. 10.1016/j.neuroimage.2011.12.090

Francey, S., O’Donoghue, B., Nelson, B., Graham, J., Baldwin, L., Yuen, H. P., Kerr, M., Ratheesh, A., Allott, K., Alvarez-Jimenez, M., Fornito, A., Harrigan, S., Thompson, A., Wood, S., Berk, M., & McGorry, P. (2020, In press). Psychosocial Intervention with or without Antipsychotic Medication for First Episode Psychosis: A Randomized Noninferiority Clinical Trial. Schizophrenia Bulletin Open.

Francey, S., O’Donoghue, B., Nelson, B., Graham, J., Baldwin, L., Yuen, H. P., Kerr, M. J., Ratheesh, A., Allott, K., Alvarez-Jimenez, M., Fornito, A., Harrigan, S., Thompson, A. D., Wood, S., Berk, M., & McGorry, P. D. (2020, January 1, 2020). Psychosocial Intervention With or Without Antipsychotic Medication for First-Episode Psychosis: A Randomized Noninferiority Clinical Trial. Schizophrenia Bulletin Open, 1(sgaa015). 10.1093/schizbullopen/sgaa015

Friston, K. J. (1998, Mar 10). The disconnection hypothesis. Schizophr Res, 30(2), 115–125. 10.1016/s0920-9964(97)00140-0

Henry, L. P., Amminger, G. P., Harris, M. G., Yuen, H. P., Harrigan, S. M., Prosser, A. L., Schwartz, O. S., Farrelly, S. E., Herrman, H., Jackson, H. J., & McGorry, P. D. (2010, 2010-06-15). The EPPIC Follow-Up Study of First-Episode Psychosis: Longer-Term Clinical and Functional Outcome 7 Years After Index Admission. The Journal of Clinical Psychiatry, 71(06), 716–728. 10.4088/JCP.08m04846yel

Karlsgodt, K. H. (2020). White matter microstructure across the psychosis spectrum. Trends in Neurosciences, 43(6), 406–416.

Klauser, P., Baker, S. T., Cropley, V. L., Bousman, C., Fornito, A., Cocchi, L., Fullerton, J. M., Rasser, P., Schall, U., & Henskens, F. (2017). White matter disruptions in schizophrenia are spatially widespread and topologically converge on brain network hubs. Schizophrenia Bulletin, 43(2), 425–435.

Kubicki, M., McCarley, R. W., & Shenton, M. E. (2005). Evidence for white matter abnormalities in schizophrenia. Current opinion in psychiatry, 18(2), 121.

Leucht, S., Kane, J. M., Kissling, W., Hamann, J., Etschel, E., & Engel, R. (2005, 10/2005). Clinical implications of Brief Psychiatric Rating Scale scores. British Journal of Psychiatry, 187(4), 366–371. 10.1192/bjp.187.4.366

Levitt, J. J., Nestor, P. G., Levin, L., Pelavin, P., Lin, P., Kubicki, M., McCarley, R. W., Shenton, M. E., & Rathi, Y. (2017). Reduced structural connectivity in frontostriatal white matter tracts in the associative loop in schizophrenia. American Journal of Psychiatry, 174(11), 1102–1111.

Li, F., Lui, S., Yao, L., Ji, G.-J., Liao, W., Sweeney, J. A., & Gong, Q. (2018). Altered white matter connectivity within and between networks in antipsychotic-naive first-episode schizophrenia. Schizophrenia Bulletin, 44(2), 409–418.

Maier-Hein, K. H., Neher, P. F., Houde, J.-C., Côté, M.-A., Garyfallidis, E., Zhong, J., Chamberland, M., Yeh, F.-C., Lin, Y.-C., Ji, Q., Reddick, W. E., Glass, J. O., Chen, D. Q., Feng, Y., Gao, C., Wu, Y., Ma, J., He, R., Li, Q., Westin, C.-F., Deslauriers-Gauthier, S., González, J. O. O., Paquette, M., St-Jean, S., Girard, G., Rheault, F., Sidhu, J., Tax, C. M. W., Guo, F., Mesri, H. Y., Dávid, S., Froeling, M., Heemskerk, A. M., Leemans, A., Boré, A., Pinsard, B., Bedetti, C., Desrosiers, M., Brambati, S., Doyon, J., Sarica, A., Vasta, R., Cerasa, A., Quattrone, A., Yeatman, J., Khan, A. R., Hodges, W., Alexander, S., Romascano, D., Barakovic, M., Auría, A., Esteban, O., Lemkaddem, A., Thiran, J.-P., Cetingul, H. E., Odry, B. L., Mailhe, B., Nadar, M. S., Pizzagalli, F., Prasad, G., Villalon-Reina, J. E., Galvis, J., Thompson, P. M., Requejo, F. D. S., Laguna, P. L., Lacerda, L. M., Barrett, R., Dell’Acqua, F., Catani, M., Petit, L., Caruyer, E., Daducci, A., Dyrby, T. B., Holland-Letz, T., Hilgetag, C. C., Stieltjes, B., & Descoteaux, M. (2017, 2017/11/07). The challenge of mapping the human connectome based on diffusion tractography. Nature Communications, 8(1), 1349. 10.1038/s41467-017-01285-x

Meng, L., Li, K., Li, W., Xiao, Y., Lui, S., Sweeney, J. A., & Gong, Q. (2018, 2018-08-31). Widespread white-matter microstructure integrity reduction in first-episode schizophrenia patients after acute antipsychotic treatment. Schizophrenia Research. 10.1016/J.SCHRES.2018.08.021

Mills, E. D. (2013). Adjusting for covariates in zero-inflated gamma and zero-inflated log-normal models for semicontinuous data. The University of Iowa.

Moser, D. A., Doucet, G. E., Lee, W. H., Rasgon, A., Krinsky, H., Leibu, E., Ing, A., Schumann, G., Rasgon, N., & Frangou, S. (2018). Multivariate associations among behavioral, clinical, and multimodal imaging phenotypes in patients with psychosis. JAMA Psychiatry, 75(4), 386–395.

Nelson, M. C., Royer, J., Lu, W. D., Leppert, I. R., Campbell, J. S., Schiavi, S., Jin, H., Tavakol, S., Vos de Wael, R., & Rodriguez-Cruces, R. (2023). The Human Brain Connectome Weighted by the Myelin Content and Total Intra-Axonal Cross-Sectional Area of White Matter Tracts. Network Neuroscience, 1–68.

O’Donoghue, B., Francey, S. M., Nelson, B., Ratheesh, A., Allott, K., Graham, J., Baldwin, L., Alvarez-Jimenez, M., Thompson, A., Fornito, A., Polari, A., Berk, M., Macneil, C., Crisp, K., Pantelis, C., Yuen, H. P., Harrigan, S., & McGorry, P. (2019). Staged treatment and acceptability guidelines in early psychosis study (STAGES): A randomized placebo controlled trial of intensive psychosocial treatment plus or minus antipsychotic medication for first-episode psychosis with low-risk of self-harm or aggression. Study protocol and baseline characteristics of participants. Early Intervention in Psychiatry, 0(0). 10.1111/eip.12716

Parnanzone, S., Serrone, D., Rossetti, M. C., D’Onofrio, S., Splendiani, A., Micelli, V., Rossi, A., & Pacitti, F. (2017, 2017/03/01). Alterations of cerebral white matter structure in psychosis and their clinical correlations: a systematic review of Diffusion Tensor Imaging studies. Rivista di Psichiatria, 52(2), 49–66.

Qiu, A., & Liu, C. (2023). Pathways link environmental and genetic factors with structural brain networks and psychopathology in youth. Neuropsychopharmacology, 48(7), 1042–1051.

Quan, M., Lee, S.-H., Kubicki, M., Kikinis, Z., Rathi, Y., Seidman, L. J., Mesholam-Gately, R. I., Goldstein, J. M., McCarley, R. W., & Shenton, M. E. (2013). White matter tract abnormalities between rostral middle frontal gyrus, inferior frontal gyrus and striatum in first-episode schizophrenia. Schizophrenia Research, 145(1-3), 1–10.

Raghava, J. M., Mandl, R. C., Nielsen, M. Ø., Fagerlund, B., Glenthøj, B. Y., Rostrup, E., & Ebdrup, B. H. (2021). Multimodal assessment of white matter microstructure in antipsychotic-naïve schizophrenia patients and confounding effects of recreational drug use. Brain Imaging and Behavior, 15(1), 36–48.

Rakesh, D., & Whittle, S. (2021). Socioeconomic status and the developing brain–A systematic review of neuroimaging findings in youth. Neuroscience & Biobehavioral Reviews, 130, 379–407.

Repple, J., Gruber, M., Mauritz, M., de Lange, S. C., Winter, N. R., Opel, N., Goltermann, J., Meinert, S., Grotegerd, D., & Leehr, E. J. (2023). Shared and specific patterns of structural brain connectivity across affective and psychotic disorders. Biological Psychiatry, 93(2), 178–186.

Romme, I. A., de Reus, M. A., Ophoff, R. A., Kahn, R. S., & van den Heuvel, M. P. (2017). Connectome disconnectivity and cortical gene expression in patients with schizophrenia. Biological Psychiatry, 81(6), 495–502.

Sabaroedin, K., Razi, A., Chopra, S., Tran, N., Pozaruk, A., Chen, Z., Finlay, A., Nelson, B., Allott, K., & Alvarez-Jimenez, M. (2023). Frontostriatothalamic effective connectivity and dopaminergic function in the psychosis continuum. Brain, 146(1), 372–386.

Sabaroedin, K., Tiego, J., & Fornito, A. (2022). Circuit-based approaches to understanding corticostriatothalamic dysfunction across the psychosis continuum. Biological Psychiatry.

Sabaroedin, K., Tiego, J., Parkes, L., Sforazzini, F., Finlay, A., Johnson, B., Pinar, A., Cropley, V., Harrison, B. J., Zalesky, A., Pantelis, C., Bellgrove, M., & Fornito, A. (2019, 07/2019). Functional Connectivity of Corticostriatal Circuitry and Psychosis-like Experiences in the General Community. Biological Psychiatry, 86(1), 16–24. 10.1016/j.biopsych.2019.02.013

Sarpal, D. K., Argyelan, M., Robinson, D. G., Szeszko, P. R., Karlsgodt, K. H., John, M., Weissman, N., Gallego, J. A., Kane, J. M., & Lencz, T. (2016). Baseline striatal functional connectivity as a predictor of response to antipsychotic drug treatment. American Journal of Psychiatry, 173(1), 69–77.

Sarpal, D. K., Robinson, D. G., Lencz, T., Argyelan, M., Ikuta, T., Karlsgodt, K., Gallego, J. A., Kane, J. M., Szeszko, P. R., & Malhotra, A. K. (2015, 2015-1-1). Antipsychotic Treatment and Functional Connectivity of the Striatum: a Prospective Controlled Study in First-Episode Schizophrenia. JAMA Psychiatry, 72(1), 5–13. 10.1001/jamapsychiatry.2014.1734

Schaefer, A., Kong, R., Gordon, E. M., Laumann, T. O., Zuo, X.-N., Holmes, A. J., Eickhoff, S. B., & Yeo, B. T. T. (2018, 09 01, 2018). Local-Global Parcellation of the Human Cerebral Cortex from Intrinsic Functional Connectivity MRI. Cerebral Cortex (New York, N.Y.: 1991), 28(9), 3095–3114. 10.1093/cercor/bhx179

Schiavi, S., Ocampo-Pineda, M., Barakovic, M., Petit, L., Descoteaux, M., Thiran, J.-P., & Daducci, A. (2020). A new method for accurate in vivo mapping of human brain connections using microstructural and anatomical information. Science Advances, 6(31), eaba8245–eaba8245. 10.1126/sciadv.aba8245

Schilling, K. G., Nath, V., Hansen, C., Parvathaneni, P., Blaber, J., Gao, Y., Neher, P., Aydogan, D. B., Shi, Y., Ocampo-Pineda, M., Schiavi, S., Daducci, A., Girard, G., Barakovic, M., Rafael-Patino, J., Romascano, D., Rensonnet, G., Pizzolato, M., Bates, A., Fischi, E., Thiran, J. P., Canales-Rodríguez, E. J., Huang, C., Zhu, H., Zhong, L., Cabeen, R., Toga, A. W., Rheault, F., Theaud, G., Houde, J. C., Sidhu, J., Chamberland, M., Westin, C. F., Dyrby, T. B., Verma, R., Rathi, Y., Irfanoglu, M. O., Thomas, C., Pierpaoli, C., Descoteaux, M., Anderson, A. W., & Landman, B. A. (2019, Jan 15). Limits to anatomical accuracy of diffusion tractography using modern approaches. Neuroimage, 185, 1–11. 10.1016/j.neuroimage.2018.10.029

Shen, X., Finn, E. S., Scheinost, D., Rosenberg, M. D., Chun, M. M., Papademetris, X., & Constable, R. T. (2017). Using connectome-based predictive modeling to predict individual behavior from brain connectivity. Nature protocols, 12(3), 506–518.

Sporns, O. (2011). The human connectome: a complex network. Annals of the new York Academy of Sciences, 1224(1), 109–125.

Stephan, K. E., Friston, K. J., & Frith, C. D. (2009, May). Dysconnection in schizophrenia: from abnormal synaptic plasticity to failures of self-monitoring. Schizophr Bull, 35(3), 509–527. 10.1093/schbul/sbn176

Szeszko, P. R., Robinson, D. G., Ikuta, T., Peters, B. D., Gallego, J. A., Kane, J., & Malhotra, A. K. (2014, May). White matter changes associated with antipsychotic treatment in first-episode psychosis. Neuropsychopharmacology, 39(6), 1324–1331. 10.1038/npp.2013.288

Theaud, G., Houde, J.-C., Boré, A., Rheault, F., Morency, F., & Descoteaux, M. (2020, 2020/09/01/). TractoFlow: A robust, efficient and reproducible diffusion MRI pipeline leveraging Nextflow & Singularity. Neuroimage, 218, 116889. 10.1016/j.neuroimage.2020.116889

Tian, Y., Margulies, D. S., Breakspear, M., & Zalesky, A. (2020, 2020-11). Topographic organization of the human subcortex unveiled with functional connectivity gradients. Nature Neuroscience, 23(11), 1421–1432. 10.1038/s41593-020-00711-6

Trépanier, M. O., Hopperton, K. E., Mizrahi, R., Mechawar, N., & Bazinet, R. P. (2016, 2016/08/01). Postmortem evidence of cerebral inflammation in schizophrenia: a systematic review. Molecular Psychiatry, 21(8), 1009–1026. 10.1038/mp.2016.90

Van Den Heuvel, M. P., Scholtens, L. H., De Lange, S. C., Pijnenburg, R., Cahn, W., Van Haren, N. E., Sommer, I. E., Bozzali, M., Koch, K., & Boks, M. P. (2019). Evolutionary modifications in human brain connectivity associated with schizophrenia. Brain, 142(12), 3991–4002.

Voineskos, A. N., Lobaugh, N. J., Bouix, S., Rajji, T. K., Miranda, D., Kennedy, J. L., Mulsant, B. H., Pollock, B. G., & Shenton, M. E. (2010). Diffusion tensor tractography findings in schizophrenia across the adult lifespan. Brain, 133(5), 1494–1504.

Voineskos, A. N., Mulsant, B. H., Dickie, E. W., Neufeld, N. H., Rothschild, A. J., Whyte, E. M., Meyers, B. S., Alexopoulos, G. S., Hoptman, M. J., Lerch, J. P., & Flint, A. J. (2020, 2020/02/26). Effects of Antipsychotic Medication on Brain Structure in Patients With Major Depressive Disorder and Psychotic Features: Neuroimaging Findings in the Context of a Randomized Placebo-Controlled Clinical Trial. JAMA Psychiatry. 10.1001/jamapsychiatry.2020.0036

Wang, Y., Wei, Y., Edmiston, E. K., Womer, F. Y., Zhang, X., Duan, J., Zhu, Y., Zhang, R., Yin, Z., & Zhang, Y. (2020). Altered structural connectivity and cytokine levels in Schizophrenia and Genetic high-risk individuals: Associations with disease states and vulnerability. Schizophrenia Research, 223, 158–165.

Wernicke, C. (1906). Grundriss der Psychiatrie in klinischen Vorlesungen. Thieme.

Wheeler, A. L., & Voineskos, A. N. (2014, 2014-08-25). A review of structural neuroimaging in schizophrenia: from connectivity to connectomics. Frontiers in Human Neuroscience, 8. 10.3389/fnhum.2014.00653

Winkler, A. M., Ridgway, G. R., Webster, M. A., Smith, S. M., & Nichols, T. E. (2014, 2014-05-15). Permutation inference for the general linear model. Neuroimage, 92, 381–397. 10.1016/J.NEUROIMAGE.2014.01.060

Yeo, B. T., Krienen, F. M., Sepulcre, J., Sabuncu, M. R., Lashkari, D., Hollinshead, M., Roffman, J. L., Smoller, J. W., Zöllei, L., & Polimeni, J. R. (2011). The organization of the human cerebral cortex estimated by intrinsic functional connectivity. Journal of neurophysiology.

Zalesky, A., Fornito, A., & Bullmore, E. T. (2010). Network-based statistic: identifying differences in brain networks. Neuroimage, 53(4), 1197–1207.

Zalesky, A., Fornito, A., Seal, M. L., Cocchi, L., Westin, C.-F., Bullmore, E. T., Egan, G. F., & Pantelis, C. (2011). Disrupted axonal fiber connectivity in schizophrenia. Biological Psychiatry, 69(1), 80–89.

Zhang, R., Wei, Q., Kang, Z., Zalesky, A., Li, M., Xu, Y., Li, L., Wang, J., Zheng, L., & Wang, B. (2015). Disrupted brain anatomical connectivity in medication-naïve patients with first-episode schizophrenia. Brain Structure and Function, 220, 1145–1159.

Zhao, W., Guo, S., Linli, Z., Yang, A. C., Lin, C.-P., & Tsai, S.-J. (2020). Functional, anatomical, and morphological networks highlight the role of basal ganglia–thalamus–cortex circuits in schizophrenia. Schizophrenia Bulletin, 46(2), 422–431.

