## Supplement for "Brain-wide Disruptions of Anatomical Connectivity in Antipsychotic-Naïve First Episode Psychosis"

Supplementary Information - MRI acquisition and processing details

Supplementary Information – Further information on the Network Based Statistic

Supplementary Table 1 – Sample characteristics

Supplementary Figure 1 – Results after excluding patients with substance induced psychosis or a missing diagnosis

Supplementary Figure 2 – Distributions of change scores

Supplementary Figure 3 – Results using alternate component forming thresholds for the NBS

Supplementary Figure 4 – Results using alternate CPM feature selection significance thresholds for significant models

Supplementary Figure 5 – Interpretations based on more conservative thresholds for connections implicated in predicting behavioural change in significant models.

Supplementary Figure 6 – Results using alternate zero-inclusion thresholds

Supplementary Figure 7 – Feature weights for model predicting longitudinal change in secondary functional outcomes scale (WHOQ)

Supplementary Figure 8 – Prediction performance of all primary and secondary models tested.

**MRI acquisition and processing details**

***MRI acquisition***

Diffusion-weighted imaging (DWI) data were acquired using a 3T Siemens Trio Tim scanner with a 32-channel head coil at the Royal Children's Hospital in Melbourne, Australia; an interleaved acquisition with the following parameters 2.3 mm^3^ voxel size, TR=7750, TE=112, FOV 240mm, 60 directions with b=3000 s/mm^2^ and 10 b=0 s/mm^2^ volumes. Structural T1-weighted (T1w; MPRAGE) scans were acquired with acquisition parameters as follow: 176 sagittal slices, with a 1mm^3^ voxel size, bandwidth 236 Hz/pixel, field of view (FOV) =256×256, matrix = 256×256×176, repetition time (TR)= 2300ms, echo time (TE) = 2.98ms and a 9° flip angle.

***MRI data processing***

During visual inspection of DWI data, 9 participants (2 controls) were excluded due to limited field-of-view. The remaining DWI data were first denoised using the *dwidenoise* tool from *MRtrix3* and then skull-stripped using *FSL* *bet*. We next implemented the synthesized b0 for diffusion distortion correction method (Synb0-DisCo; Schilling et al., 2019), which is a validated method that uses T1w images to create a synthetic undistorted b0 image for susceptibility and distortion correction. Diffusion data were corrected for susceptibility and signal outliers, eddy-current-induced distortions, slice dropouts, gradient-non-linearities, subject motion and susceptibility-by-motion (Andersson et al., 2017; Andersson et al., 2016). We then implemented the *tractoflow* pipeline (Theaud et al., 2020), which included the following steps. First, the DWI data were bias-corrected using the N4 tool as implemented in *Advanced Normalization Tools* (ANTs; Avants et al., 2011). The image was then cropped using *Dipy* (Garyfallidis et al., 2014) and normalized to a mean image value of approximately 1000 using *dwinormalise* from *MRtrix3* (Tournier et al., 2019). The data were then resampled to 1 mm isotropic spatial resolution and the *Dipy* *TensorModel* was used to estimate the Diffusion Tensor Image (Basser et al., 1994) at every voxel, with a weighted least squares method. The *csdeconv* package from *Dipy* was used to compute fibre orientation distributions (FOD), which represents the estimated orientation distribution of fibre structure at each voxel (Descoteaux et al., 2009; Tournier et al., 2007). Peaks representing primary diffusion directions were extracted from local maxima of each FOD’s angular distribution. This FOD field was later used for tractography and to estimate structural connectivity.

The T1w data were processed using the same protocol as DWI data for denoising, N4 bias correction, resampling, brain extraction, and cropping. The T1w data were then registered to the b0 image using non-linear ANTs (Avants et al., 2011). Segmentation of grey matter, white matter, subcortex and cerebrospinal fluid was performed using *CIVIT* (Ad-Dab’bagh et al., 2006), and the resulting tissue partial volume estimate maps were used to compute the inclusion and exclusion masks for where streamlines can traverse, as well as a grey/white matter interface mask used for seeding (Theaud et al., 2020).

Probabilistic tractography was preformed using a particle filtering tractography algorithm (Girard et al., 2014) implemented in Dipy. Similar to anatomically constrained tractography (Smith et al., 2012), particle filtering tractography takes advantage of previously computed tissue maps to define areas where streamline can traverse. The white matter/grey matter interface was used for seeding 15,000,000 streamlines for each subject. Default parameters were used for other tracking options (Al-Sharif et al., 2020).

Importantly, probabilistic tractography algorithms are prone to false positives and do not directly index the quantitative strength of connections between pairs of regions (Maier-Hein et al., 2017; Schilling et al., 2019). We therefore implemented a state-of-the-art optimisation procedure, Convex Optimization Modelling for Microstructure Informed Tractography (COMMIT2), which is superior to other methods on key benchmarks derived from fibre-tracking phantoms (Nelson et al., 2023; Schiavi et al., 2020). COMMIT2 uses a forward model to recover the connectome with the minimum number of bundles that best explains the local axon density estimated from the DWI signal (Schiavi et al., 2020). In doing so, COMMIT2 filters and re-weights pair-wise connections strengths according to their contribution to the observed DWI signal, providing more biologically accurate and quantitative estimates of connectivity than traditional streamline count measures.

To create a whole-brain structural connectivity matrix, COMMIT2-re-weighted streamlines were assigned to the closest region in a previously validated combined 300-region cortical (Schaefer et al., 2018) and 32-region subcortical (Tian et al., 2020) atlas, within a 2-mm radius of the streamline endpoints (Tournier et al., 2019). Thirteen somatomotor and temporal pole regions were excluded due to limited field-of-view within the DWI data, yielding undirected structural connectivity matrices defining pair-wise connections for 319 parcellated regions.

**Further information on zero-inflated gamma regression**

The zero-inflated gamma regression model is a two-part hurdle model comprising a zero-inflation component and a gamma component. The zero-inflation component is modelled using a logistic regression and accounts for the difference in the probability of observing a zero at a given edge. The gamma component uses logarithmic link function and characterizes differences in positive non-zero connectivity strength at a given edge.

For a given edge, if no participant had a zero value, or the log-regression component of the model failed to converge due to an insufficient number of zeros, only the gamma regression component of the model was evaluated. Neither part of the zero-inflated model is likely to converge when the data almost entirely consists of zeros, so we excluded edges with >80% zero-values. We then repeated our primary analyses at thresholds of 70% and 90% to ensure that our results were robust to this parameter choice (see SFig6). Model diagnostics were assessed at each edge using the Distributed Hierarchical Modelling and Analysis for Robust Inference package (DHARMa; Hartig, 2020). This package tests for dispersion (when the variability in connectivity strength is lower/higher than expected based on the gamma distribution), heteroscedasticity, and outliers using a simulation-based approach to create readily interpretable residuals for generalized zero-inflated models (for details see Hartig, 2020). Any edge where the model was significantly ($p<.05$) over/under-dispersed, heteroscedastic, or with substantial outliers was excluded from further analysis (4.47% of edges were excluded based on these model diagnostics).

All continuous covariates were centred. Out of a total of 50,721 possible pairwise connections between 319 brain regions, 21,476 edges were included in the final analysis. At each included edge, the Wald test statistic was computed and used to derive a parametric *p*-value for each of the two-parts of the zero-inflated gamma regression model, with the minimum $p$-value between the two models considered for further analysis (Lee et al., 2012). We repeated the above procedure after permuting the group assignment between patients and controls 10,000 times, leading to the generation of 10,000 null matrices of Wald test statistics and corresponding $p$-values.

**Further information on the Network Based Statistic**

The Network Based Statistic (NBS) was used to perform family-wise error-corrected (FWE) inference at the level of connected-components of edges showing a common effect, resulting in a substantial boost in statistical power compared to mass univariate analysis (Zalesky et al., 2010). Connected components refer to sets of supra-threshold edges that can be linked by a path. The NBS procedure involves setting a primary component-forming threshold, $\tau,$ to both the observed and permuted data. The choice of this threshold is arbitrary; more lenient thresholds will be sensitive to weaker differences distributed over a large number of edges while more stringent thresholds will be sensitive to stronger effects possibly extending over smaller subsets of edges. We report here results for $\tau$ set to $p<.05$ and show results for $p<.01$and $p<.001$ are presented in SFig 3. For both the observed and permuted null data, the size (number of edges) of the connected components in the supra-thresholded network was recorded. The size of largest component from each permutation was used to build a null distribution of the maximal statistic, which can be used to obtain a FWE $p$-value for each observed component as the proportion of null component sizes larger than the observed value.

Supplementary Table 1 (previously reported Chopra, Fornito, et al. (2021)) – Sample characteristics

|  | First Episode Psychosis (N=61) |  | Healthy control (N=27) | T / χ^2^ (p) ^†^ |
| --- | --- | --- | --- | --- |
| Baseline age, years (SD) | 19.15 (2.83) |  | 21.9 (1.93) | -4.49 (<0.001) |
| Females, N (%) | 27 (44.3%) |  | 17 (62.9%) | 2.19 (0.139) |
| Handedness, Left , N (%) | 4 (6.5%) |  | 3 (11.1%) | 0.465 (0.495) |
| Education, years (SD)  Diagnosis, N  Major depression with psychosis  Schizophreniform disorder  Psychotic disorder NOS  Substance-induced psychotic disorder  Delusional disorder    Schizophrenia  Missing diagnosis | 12.25 (2.08)  13  10  15  7  5  10  1 |  | 15.2 (1.90)  -  -  -  -  -  -  - | -6.21 (0.001) |
| Baseline BPRS Total, mean (SD) | 57.6 (9.87) |  | - |  |
| Baseline SOFAS, mean (SD) | 52.3 (12.3) |  | - |  |
| Baseline SANS, mean (SD) | 35.1 (17.65) |  | - |  |
| Baseline HAM-D, mean (SD) | 18.9 (6.67) |  | - |  |
| Baseline HAM-A, mean (SD) | 21.2 (6.8) |  | - |  |
| Baseline QLS, mean (SD) | 69.6 (22.4) |  | - |  |

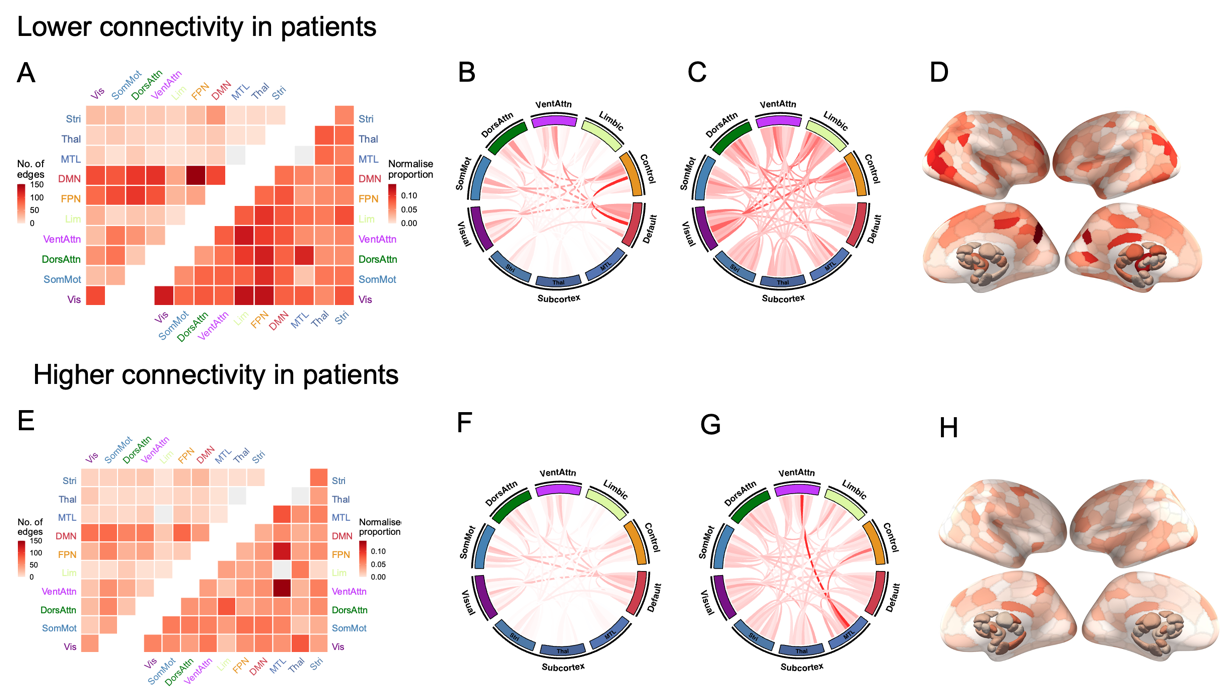

**Supplementary Figure 1** – Results examining group differences in structural connectivity after excluding subjects missing a diagnosis or substance-induced psychosis ($p_{FWE} = .02;3157 edges; \tau=0.05)$. **A/E)** heatmap of the proportion of edges within the NBS component that fall within each of the canonical networks as represented quantified using raw (upper triangle) and normalized proportions (lower triangle; see Methods for details); **B-C/F-G)** a circle plot of the proportion of edges within the NBS component that fall within each of the canonical networks as represented quantified using raw (**B/F**) and normalized **(C/G**) proportion; **D/H**) surface renderings depicting the number of edges in the NBS subnetwork attached to each brain region.

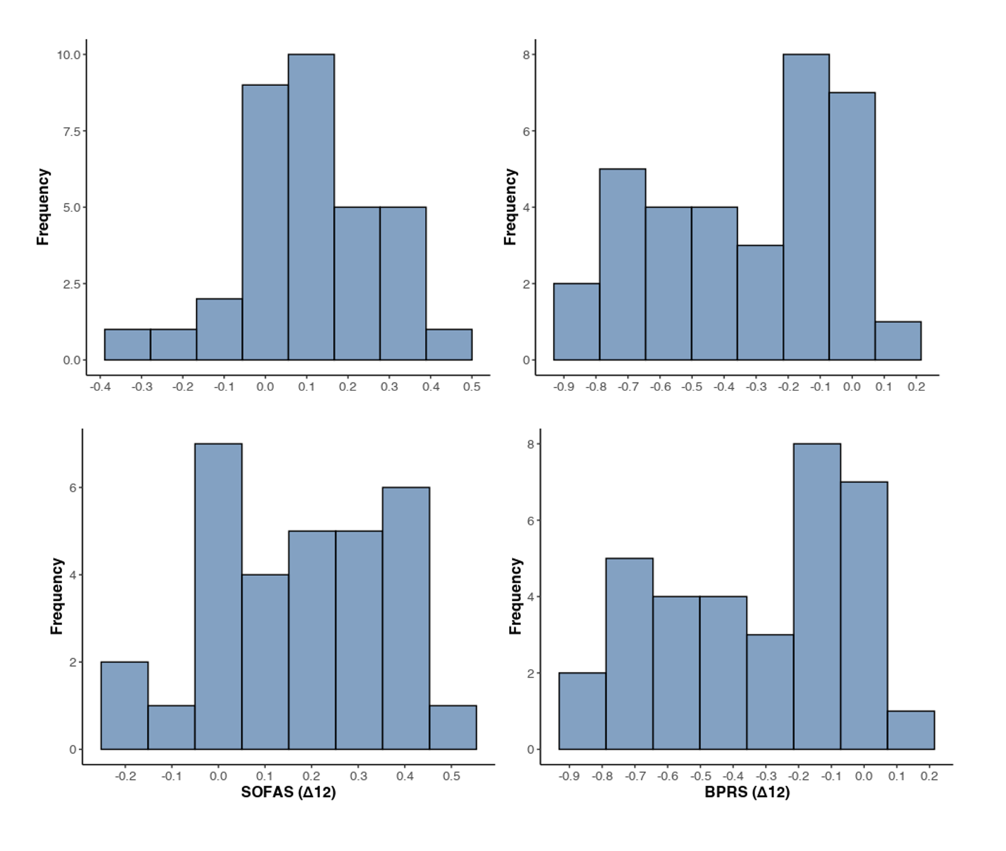

Supplementary Figure 2 – Distributions of change scores for the primary outcome measures (SOFAS and BPRS) at baseline to 3 months (Δ3) and baseline to 12 months (Δ12). Higher scores represent better outcome in SOFAS and worse outcomes in BPRS.

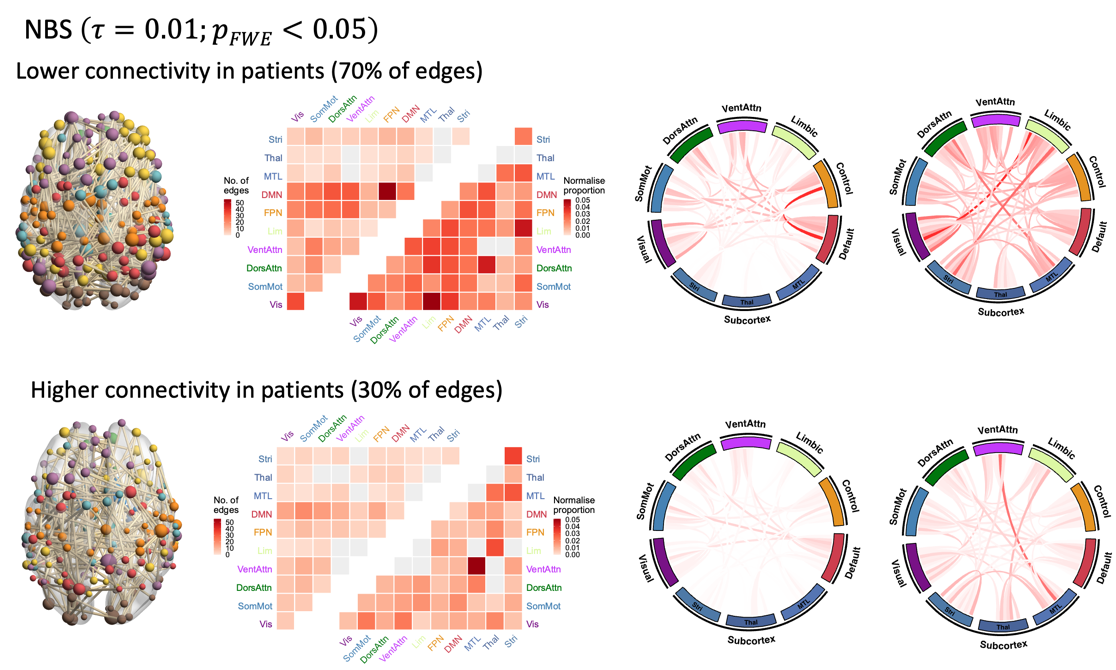

**Supplementary Figure 3** - Results using an alternate component forming threshold ($\tau<0.01)$. No significant component was detected at $\tau<0.001$, suggesting that the detected effect is diffuse and spatially widespread.

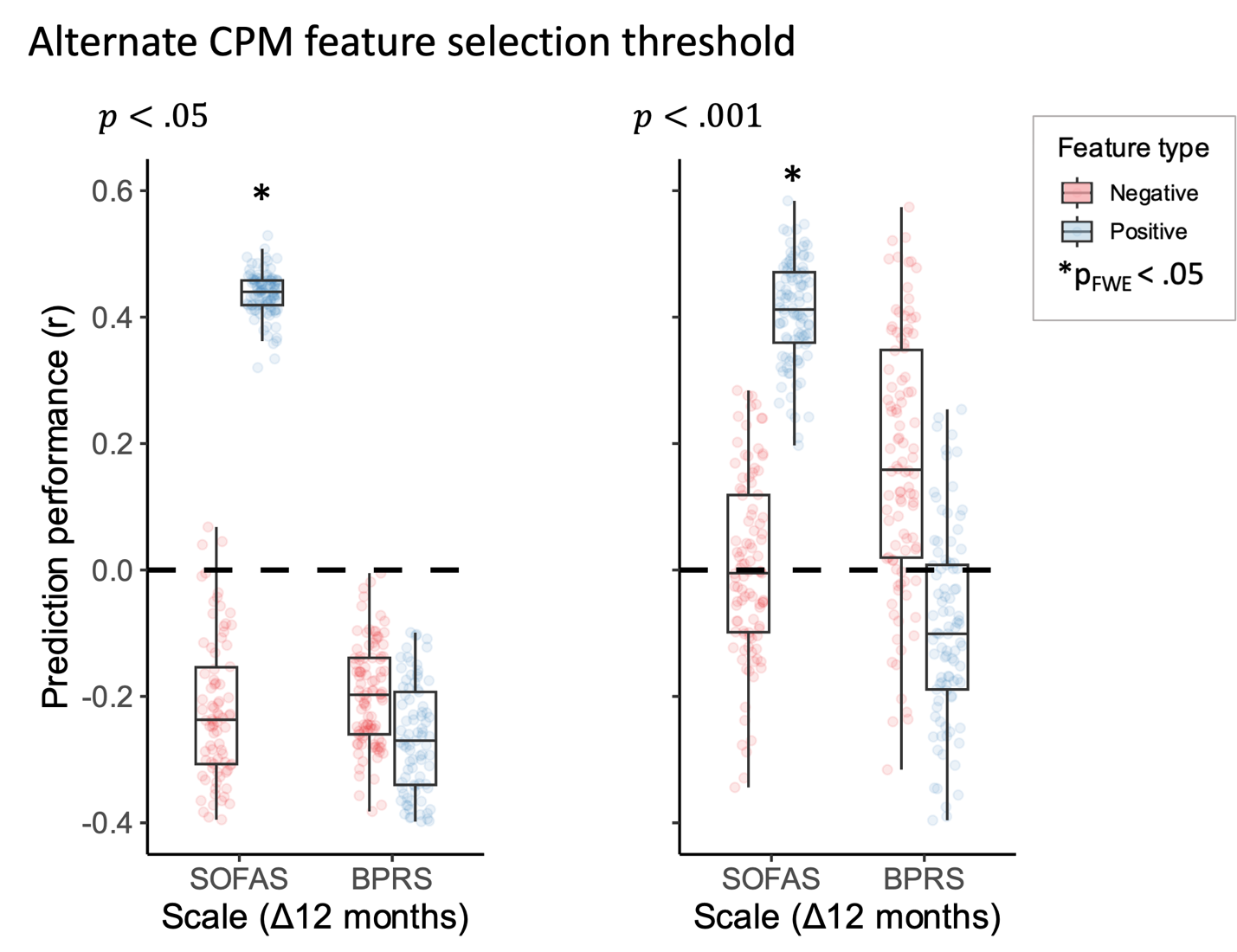

**Supplementary Figure 4** – Results for significant prediction models using alternate feature selection thresholds (p<0.05 and p<0.001).

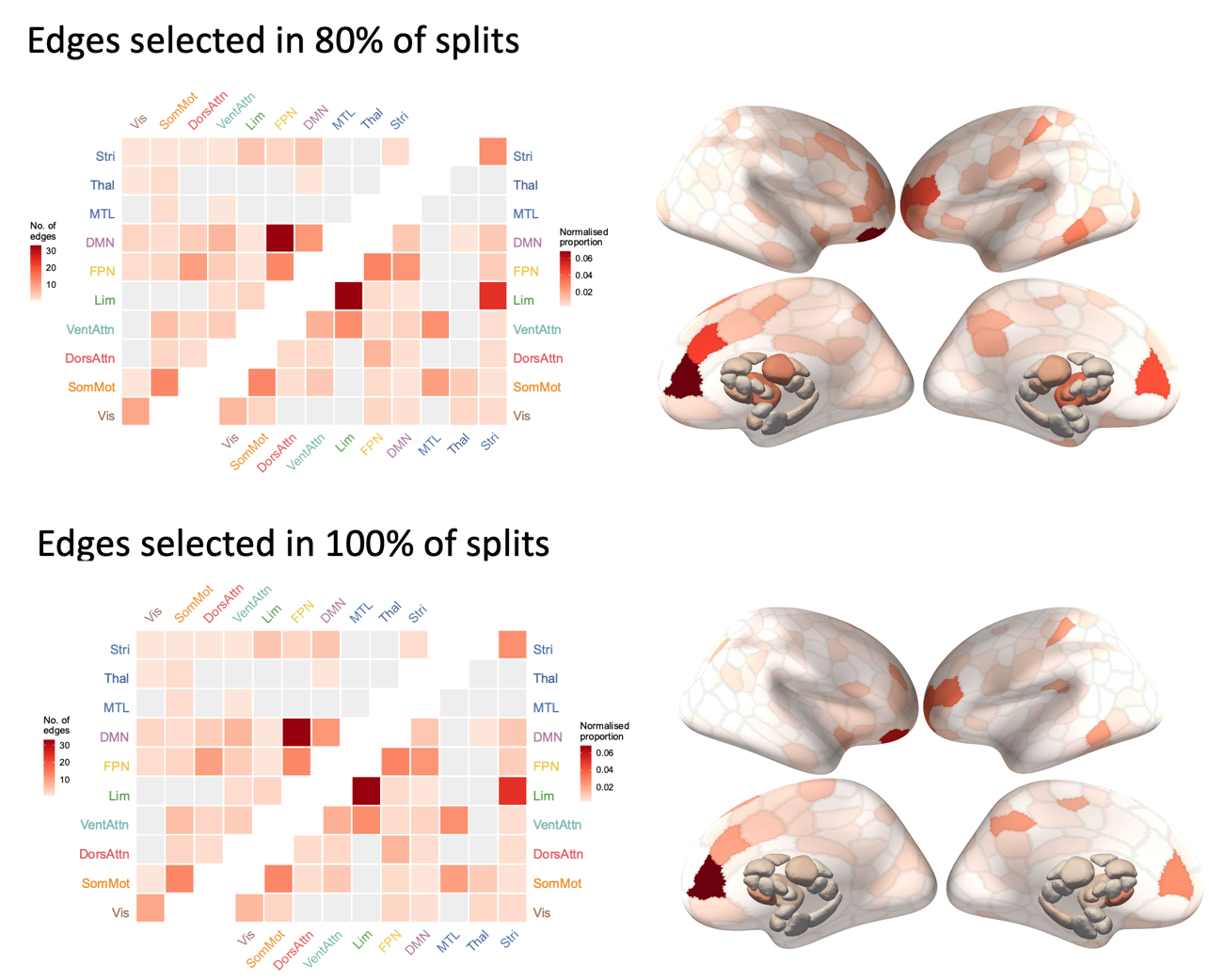

**Supplementary Figure 5** – Connections implicated in prediction of change in patient functional outcomes (SOFAS) at 12-months, based on more conservative thresholds (i.e., edges included in 80% or 100% of the 100 iterations of train/test splits.

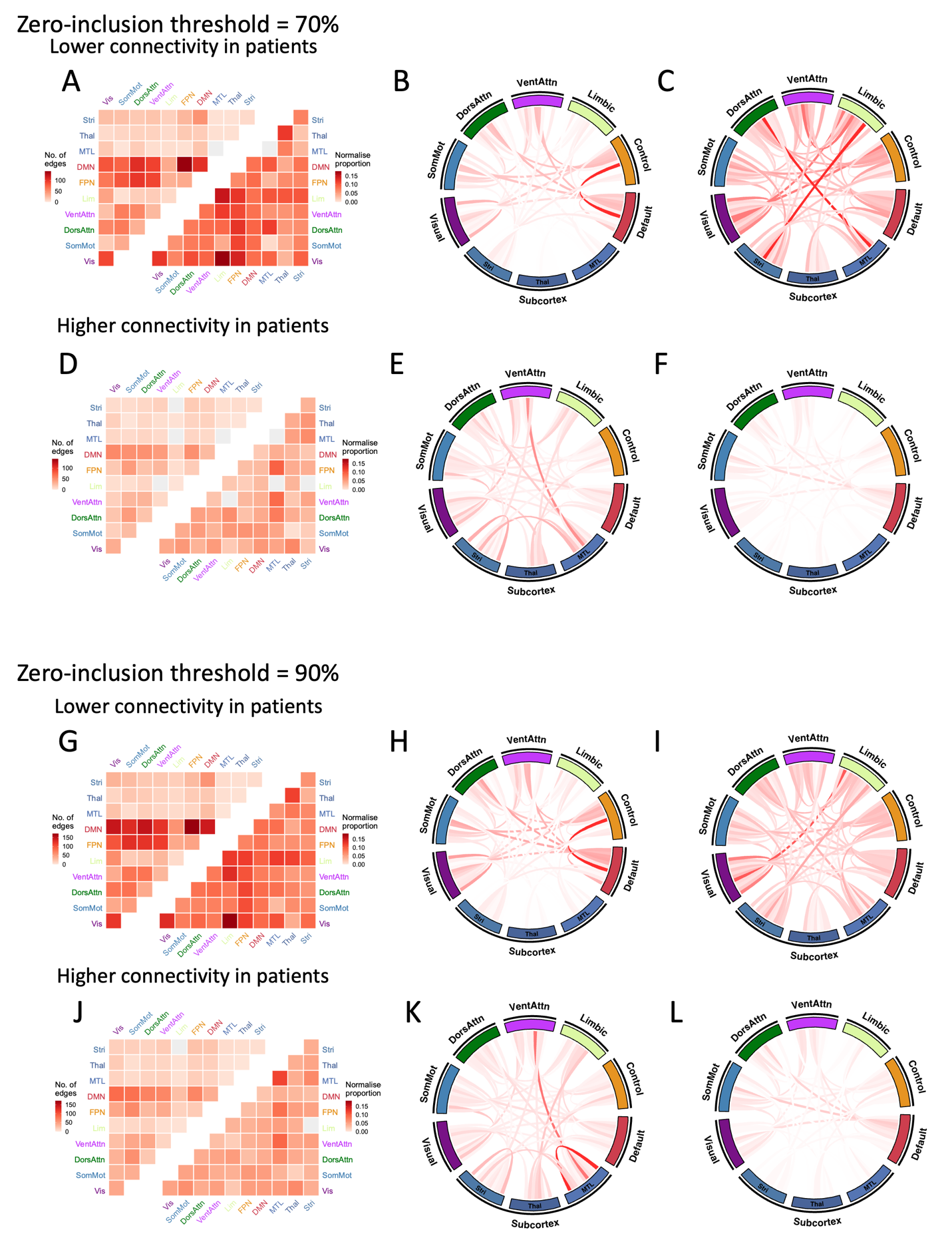

**Supplementary Figure 6**– Results examining group differences in structural connectivity using alternate zero-inclusion threshold (70%/90%; See supplement – Zero inflated gamma regression)

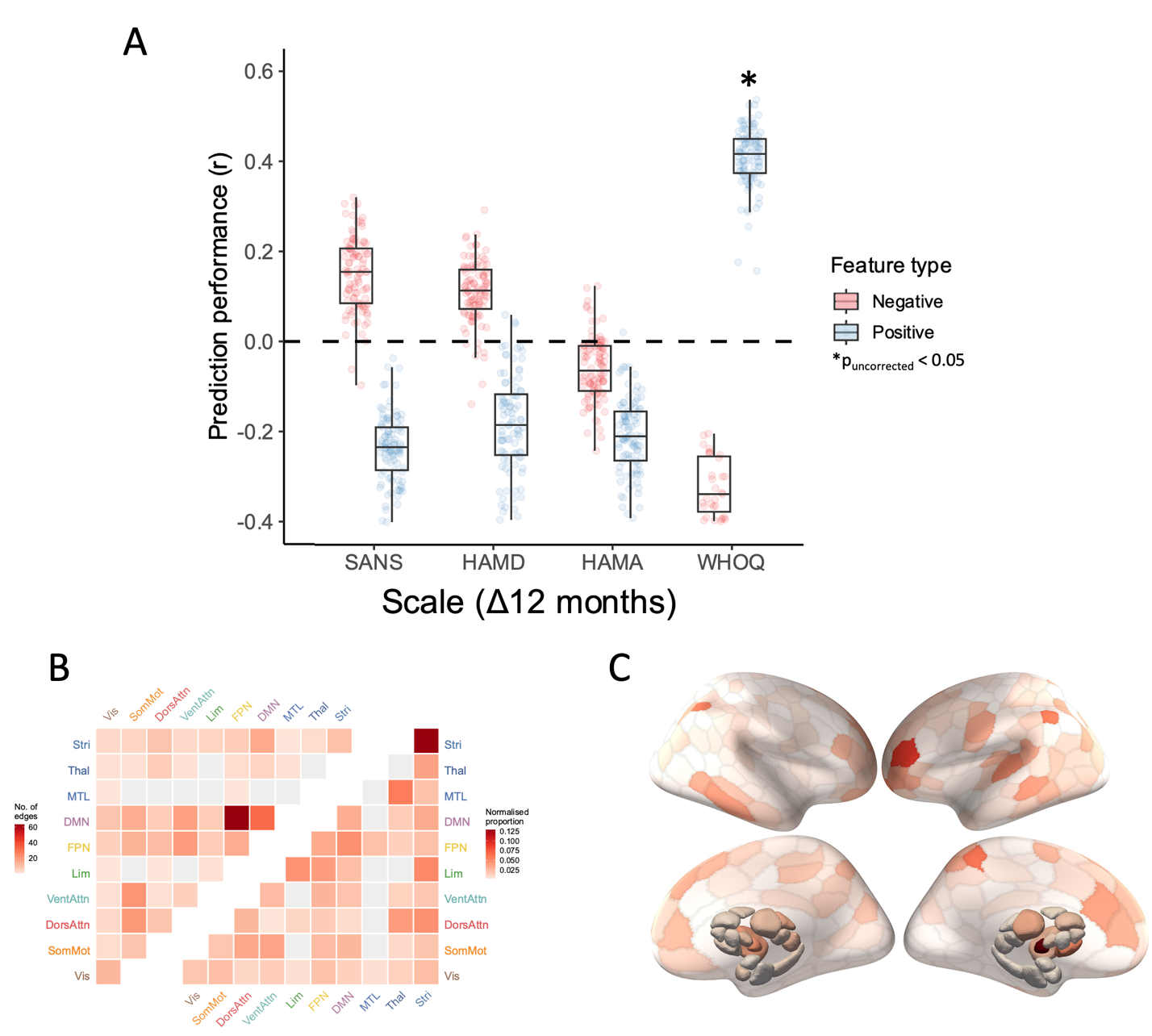

**Supplementary Figure 7** – Feature weights for models predicting longitudinal change in secondary functional outcomes scale (WHOQ).

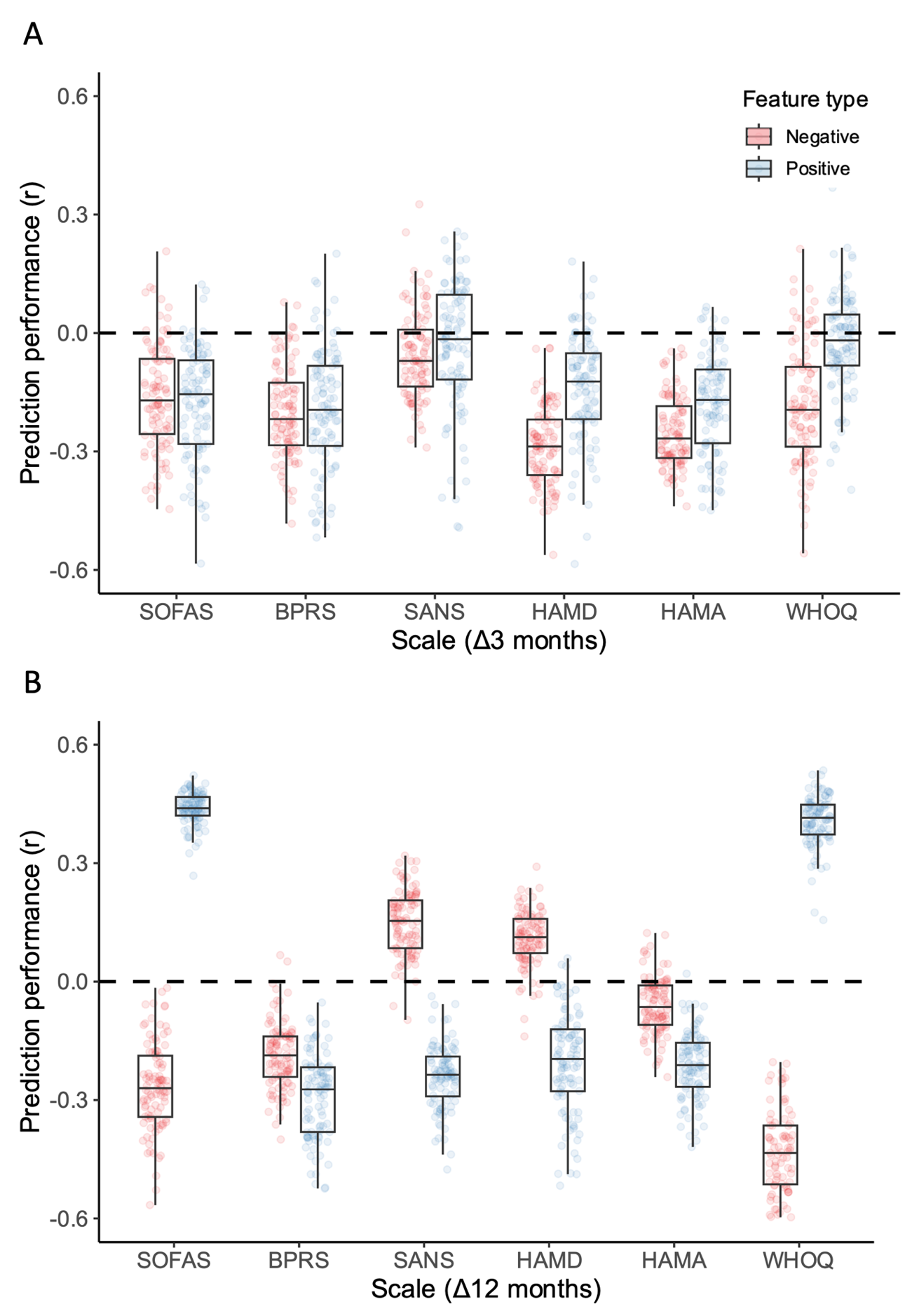

**Supplementary Figure 8** – Prediction performance of all primary and exploratory models tested.
